# Microstructural changes in the inferior tuberal hypothalamus correlate with daytime sleepiness in Lewy body disease

**DOI:** 10.1101/2024.08.16.24312102

**Authors:** Jesse S. Cohen, Hamsanandini Radhakrishnan, Christopher A. Olm, Sandhitsu R. Das, Philip A. Cook, David A. Wolk, Daniel Weintraub, David J. Irwin, Corey T. McMillan

## Abstract

**Background:** Excessive daytime sleepiness (EDS) is a disabling symptom of Lewy body disorders (LBD). The hypothalamus is a key sleep-wake regulator, but its contribution to EDS in LBD remains unclear.

**Objectives:** Use diffusion MRI to evaluate the relationship of hypothalamic microstructure to EDS symptoms in LBD.

**Methods:** We studied 102 patients with clinically-defined LBD (Parkinson’s disease, n=93; Parkinson’s disease dementia, n=4; and dementia with Lewy bodies, n=5) and Epworth Sleepiness Scale (ESS) within 2 years of MRI. Mean diffusivity (MD) was compared between EDS+ (ESS≥10, n=37) and EDS- (ESS<10, n=65) groups in the whole hypothalamus and three subregions, covarying for age and sex. Secondary analyses tested correlations between subregion MD and continuous ESS, global cognition, and motor scores; and between subregion volume and continuous ESS.

**Results:** MD was increased in EDS+ compared to EDS-only in the inferior tuberal subregion (Cohen’s d=0.43, p=0.043, β=0.117±0.057), with trend level differences in the whole hypothalamus (Cohen’s d=0.39, p=0.064, β=0.070±0.037) and superior tuberal subregion (Cohen’s d=0.38, p=0.073, β=0.063±0.035). No difference was seen in the posterior subregion (Cohen’s d=0.1, p=0.628, β=0.019±0.038). Significant correlations with continuous ESS were seen in MD of whole hypothalamus (r^2^=0.074, p=0.0057), superior tuberal (r^2^=0.081, p=0.0038), and inferior tuberal (r^2^=0.073, p=0.0059) subregions. There was no correlation of hypothalamic MD with global cognition or motor scores, and no correlation of whole/subregional hypothalamic volumes with ESS.

**Conclusions:** Daytime sleepiness associates with increased MD in the inferior tuberal hypothalamus in an LBD cohort. This suggests degeneration within this region could contribute to EDS symptoms.

## Introduction

Sleep disorders are increasingly recognized as both cause and consequence of neurodegeneration^1^. Excessive daytime sleepiness (EDS) afflicts up to 80% of individuals with Lewy body disorders (LBD)^2-5^. LBD include Parkinson’s disease (PD), PD dementia (PDD), and Dementia with Lewy bodies (DLB), and together they are the second leading cause of neurodegenerative dementia^6^. Furthermore, EDS in LBD is associated with increased cognitive impairment^7,8^, psychiatric symptoms and institutionalization^9,10^.

Despite its significance, the mechanisms underlying EDS in LBD remain poorly understood, limiting the development of effective therapies. The hypothalamus, a central regulator of the sleep-wake cycle, has been long implicated in the pathology of LBD^11-15^. The hypothalamic tuberomammillary nucleus (TMN) is specifically affected by synuclein pathology by Parkinson disease Braak stage 3^16^. The TMN is crucial for maintaining wakefulness via production of histamine, a wake-promoting neurotransmitter^17^; antihistamines induce sleep via antagonism of TMN signaling. Thus, dysfunction of the histaminergic system in LBD could contribute to daytime sleepiness.

Previous studies of hypothalamic degeneration in LBD used structural MRI to measure volumetric differences, with inconsistent results^18-21^. While one small study found decreased hypothalamic volume in PD^19^, this was refuted by a larger study^20^ which found no difference between PD and controls.

While structural MRI evaluates brain volume (macrostructure), diffusion MRI analyzes brain microstructure by measuring the diffusion properties of water molecules within the tissue^22^. This enables interrogation of microstructural integrity not possible with conventional volumetric analyses^23^. Mean diffusivity (MD) is increased in the presence of neuronal loss and gliosis^24^, and thus may be more sensitive than structural MRI to early neurodegeneration^25,26^. In this study, we use diffusion MRI to test the relationship between severity of daytime sleepiness and hypothalamic MD in individuals with LBD.

## Methods

### Study Design

This investigation was a cohort study using data from the University of Pennsylvania Integrated Neurodegenerative Disease Database (INDD)^27,28^. All procedures were performed with informed consent under institutional review board approval.

### Study Setting

INDD combines data from several neurology clinical centers at the University of Pennsylvania from 2005-present, including the University of Pennsylvania’s Frontotemporal Degeneration Center, Parkinson’s Disease and Movement Disorders Clinic, the Philadelphia VA Parkinson’s Disease Research, Education and Clinical Center (PADRECC), and Alzheimer’s Disease Research Center.

### Participants

Individuals were retrospectively identified by querying the INDD as of 10/04/2022. LBD cases were selected if they had a clinical diagnosis of LBD (PD, PDD, DLB) at the visit closest to MRI acquisition, established through clinical consensus using published clinical criteria^29-33^. Inclusion also required (a) at least one structural brain MRI scanning session with both T1-weighted (T1w) and diffusion-weighted (DWI) images acquired and (b) Epworth sleepiness scale (ESS) within ±2 years of MRI. There were no exclusion criteria.

### Ascertainment of excessive daytime sleepiness

The Epworth Sleepiness Scale (ESS)^34^ was used to assess presence and severity of daytime sleepiness. The ESS asks the likelihood of falling asleep in eight different everyday situations recently. ESS has shown excellent psychometric properties in PD ^35^ and is widely used across LBD. An ESS score of 10 or greater is used to define pathologic sleepiness^36^, and this cutoff has been used previously to study EDS in LBD^37^. Our subsequent analysis uses ESS as a continuous outcome measure, which has also been previously validated^38^.

### MRI acquisition and analysis

All images were acquired on 3T Siemens scanners (Erlangen, Germany) at the University of Pennsylvania. If multiple MRIs were available for an individual, the MRI closest to ESS date was used.

#### Hypothalamic segmentation and quantification

Hypothalamic regions are segmented from the T1w image using the fully-automated deep learning method by Billot et al.^39^ via FreeSurfer^40^. This provides labeling for the following subregions (putative included nuclei in parentheses): anterior-superior (preoptic area, paraventricular nucleus (PVN)), anterior-inferior (suprachiasmatic nucleus, supraoptic nucleus (SON)), superior tuberal (dorsomedial nucleus, PVN, lateral hypothalamus), inferior tuberal (infundibular nucleus, ventromedial nucleus, SON, lateral tuberal nucleus, tuberomammillary nucleus (TMN)), and posterior (mammillary body, lateral hypothalamus, TMN)^39^. The anterior subregions were excluded from subsequent analysis given their small size (often <10 voxels), and since either the anterior-superior or anterior-inferior subregion was missing in some individuals. Diffusion images were preprocessed using QSI-Prep 0.18.0^41^. Scalar maps of fractional anisotropy (FA) and mean diffusivity (MD) were generated, using the FA map to register T1w and diffusion images.

Given no *a priori* hypothesis regarding laterality of MD differences and to limit multiple hypothesis testing, the mean bilateral subregion MD for each subject was calculated (i.e. left subregion + right subregion / 2) and used for all subsequent analysis.

### Clinical Evaluation

To evaluate clinical characteristics of our cohort, Mini-Mental State Examination (MMSE)^42^ was used as a measure of global cognition. For individuals lacking MMSE, Montreal Cognitive Assessment (MoCA)^43^ scores were converted to MMSE scores using a conversion table established in individuals with PD^44^. Movement Disorder Society Unified Parkinson’s Disease Rating Scale (MDS-UPRDS) Part III was used to characterize motor impairment^45^. Testing data closest to the date of the MRI was used, limited to within one year of MRI (Table 1).

**Table 1:** Demographic and clinical variables among those with and without EDS.

| Participant characteristics |  |  |  |  |
| --- | --- | --- | --- | --- |
| Variable | ESS >10<br>(EDS+)<br>N = 37 (36.3%) | ESS ≤10<br>(EDS-)<br>N = 65 (63.7%) | p-value | All participants<br>N = 102 |
| Age at ESS (years), Mean (SD) | 67.2 (7.3) | 65.2 (7.9) | 0.199<sup>1</sup> | 65.9 (7.7) |
| Sex (Males), n (%) | 26 (38.8%) | 41 (61.2%) | 0.604<sup>2</sup> | 67 (65.7%) |
| Race (White), n (%) | 37 (37.0%) | 63 (63.0%) | 0.533<sup>3</sup> | 100 (98.0%) |
| Education (years), Mean (SD) | 16.3 (2.7) | 16.5 (2.2) | 0.702<sup>4</sup> | 16.4 (2.4) |
| Diagnosis, n (%) |  |  | **0.021<sup>3</sup>** |  |
| PD | 30 (32.3%) | 63 (67.7%) |  | 93 (91.2%) |
| DLB | 4 (80.0%) | 1 (20.0%) |  | 5 (4.9%) |
| PDD | 3 (75.0%) | 1 (25.0%) |  | 4 (3.9%) |
| MMSE, Mean (SD) | 27.1 (3.7) | 28.7 (1.5) | **0.006<sup>4</sup>** | 28.2 (2.6) |
| MDS-UPDRS III, Mean (SD) | 34.3 (14.7) | 23.5 (11.4) | **<0.001<sup>4</sup>** | 27.4 (13.7) |
| Missing | 9 | 16 |  | 25 |
| LEDD (mg/day), Mean (SD) | 498.9 (308.5) | 551.7 (394.3) | 0.847<sup>4</sup> | 532.9 (365.2) |
| Missing | 2 | 2 |  | 4 |

<sup>1</sup> Welch Two Sample t-test
<sup>2</sup> Pearson's Chi-squared test
<sup>3</sup> Fisher's exact test
<sup>4</sup> Wilcoxon rank sum test
Bolded p-values are statistically significant at unadjusted $p < 0.05$ threshold. Abbreviations: ESS = Epworth sleepiness scale; EDS = excessive daytime sleepiness; SD = Standard deviation; DLB = Dementia with Lewy bodies; PDD = Parkinson's disease dementia; PD = Parkinson's disease. MMSE = Mini Mental State Exam; MDS-UPDRS III = Movement Disorder Society Unified Parkinson's Disease Rating Scale Part III. LEDD = Levodopa equivalent daily dose.

### Statistical Approach

Parametric and non-parametric tests were used for comparison of demographic variables as appropriate. Student’s t-test was used for comparisons of hypothalamic subregion MD between EDS± groups; brain measures were adjusted for age and sex. All results were considered significant at p<0.05. No adjustment for multiple comparisons was made given a limited number of prespecified hypotheses^46^. Effect sizes for differences between EDS± were calculated using Cohen’s d; suggested interpretation is 0.2 = small, 0.5 = medium, and 0.8 = large effect size^47^.

To assess the relationship between subregion MD and continuous measures of ESS, MMSE and MDS-UPDRS III scores, partial correlation was used, covarying for age and sex. Similarly, we used partial correlation to test whether hypothalamic atrophy was related to ESS to confirm MD findings were independent of atrophy.

For all statistical analyses, R version 4.2.3 was used. All statistical tests were two-sided.

### Data availability

Deidentified data supporting the findings of this study will be made available from the corresponding author upon reasonable request.

## Results

### Baseline characteristics of EDS+ vs. EDS-

Demographic and clinical characteristics are described in Table 1. There were significant differences in clinical diagnosis, MMSE, and MDS-UPDRS III scores. There were fewer individuals with a clinical phenotype of PD in the EDS+ group. The EDS+ group performed about 1.5 points worse on MMSE (lower score is worse) and about 11 points worse on MDS-UPDRS III (higher score is worse). There was no difference in proportion of males, self-reported race, education or levodopa-equivalent daily dose (LEDD) between groups.

### Hypothalamic MD in EDS+ vs. EDS-

To test whether microstructural differences in the hypothalamus arise in LBD with EDS, we compared hypothalamic MD between those with and without EDS, correcting for age and sex. MD was increased in EDS+ compared to EDS-only in the inferior tuberal subregion (Cohen’s d = 0.43, p = 0.043, β = 0.117 ± 0.057), with trend level differences in the whole hypothalamus (Cohen’s d = 0.39, p = 0.064, β = 0.070 ± 0.037) and the superior tuberal subregion (Cohen’s d = 0.38, p = 0.073, β = 0.063 ± 0.035). No difference was seen in the posterior subregion (Cohen’s d = 0.1, p = 0.628, β = 0.019 ± 0.038) (Table 2).

**Table 2:**
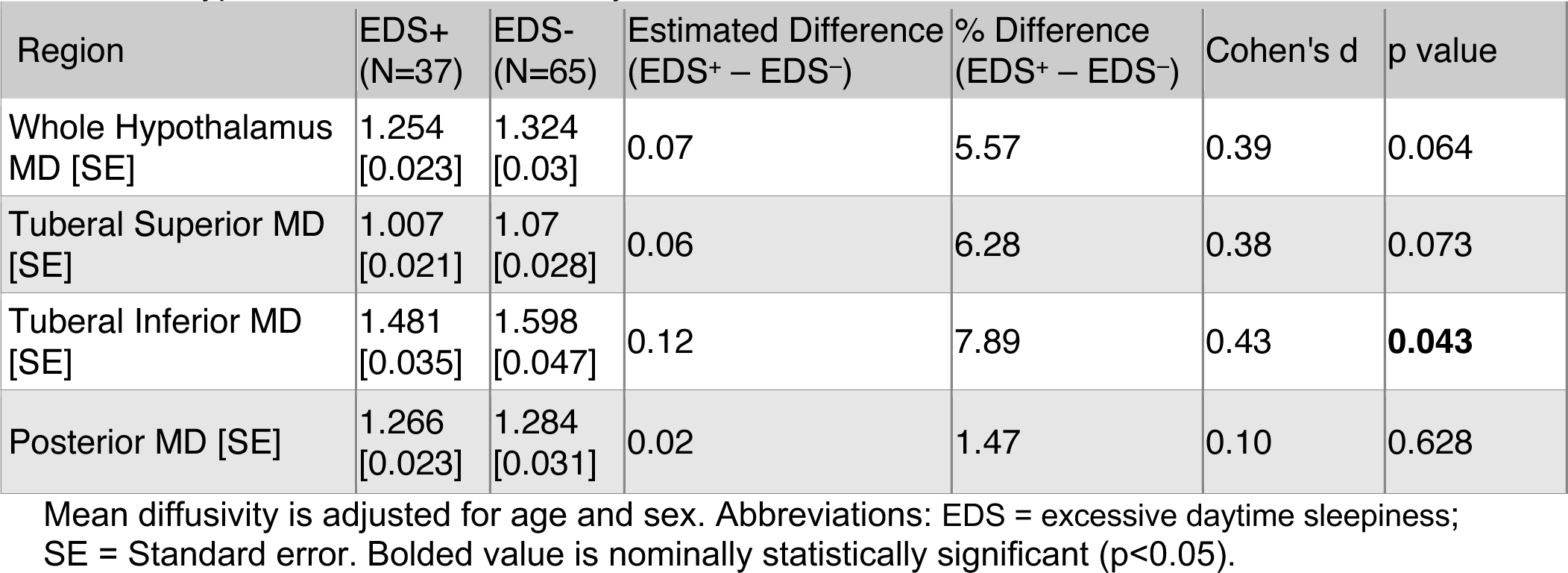
Hypothalamic mean diffusivity in EDS±.

| Region | EDS+<br>(N=37) | EDS-<br>(N=65) | Estimated Difference<br>(EDS+ – EDS-) | % Difference<br>(EDS+ – EDS-) | Cohen's d | p value |
| --- | --- | --- | --- | --- | --- | --- |
| Whole Hypothalamus MD [SE] | 1.254<br>[0.023] | 1.324<br>[0.03] | 0.07 | 5.57 | 0.39 | 0.064 |
| Tuberal Superior MD [SE] | 1.007<br>[0.021] | 1.07<br>[0.028] | 0.06 | 6.28 | 0.38 | 0.073 |
| Tuberal Inferior MD [SE] | 1.481<br>[0.035] | 1.598<br>[0.047] | 0.12 | 7.89 | 0.43 | **0.043** |
| Posterior MD [SE] | 1.266<br>[0.023] | 1.284<br>[0.031] | 0.02 | 1.47 | 0.10 | 0.628 |

Mean diffusivity is adjusted for age and sex. Abbreviations: EDS = excessive daytime sleepiness; SE = Standard error. Bolded value is nominally statistically significant ( $p < 0.05$ ).

### Correlation of hypothalamic MD with continuous sleepiness scores

To determine whether groupwise differences reflect a linear relationship between hypothalamic MD and sleepiness, we tested for correlations between hypothalamic MD and ESS, covarying for age and sex. Correlations with ESS were seen in the whole hypothalamus (R^2^ = 0.074, p = 0.0057), superior tuberal (R^2^ = 0.081, p = 0.0038), and inferior tuberal subregions (R^2^ = 0.073, p = 0059). No correlation was seen in the posterior subregion (R^2^ =0.012, p = 0.27) (Figure 1).

**Figure 1:**
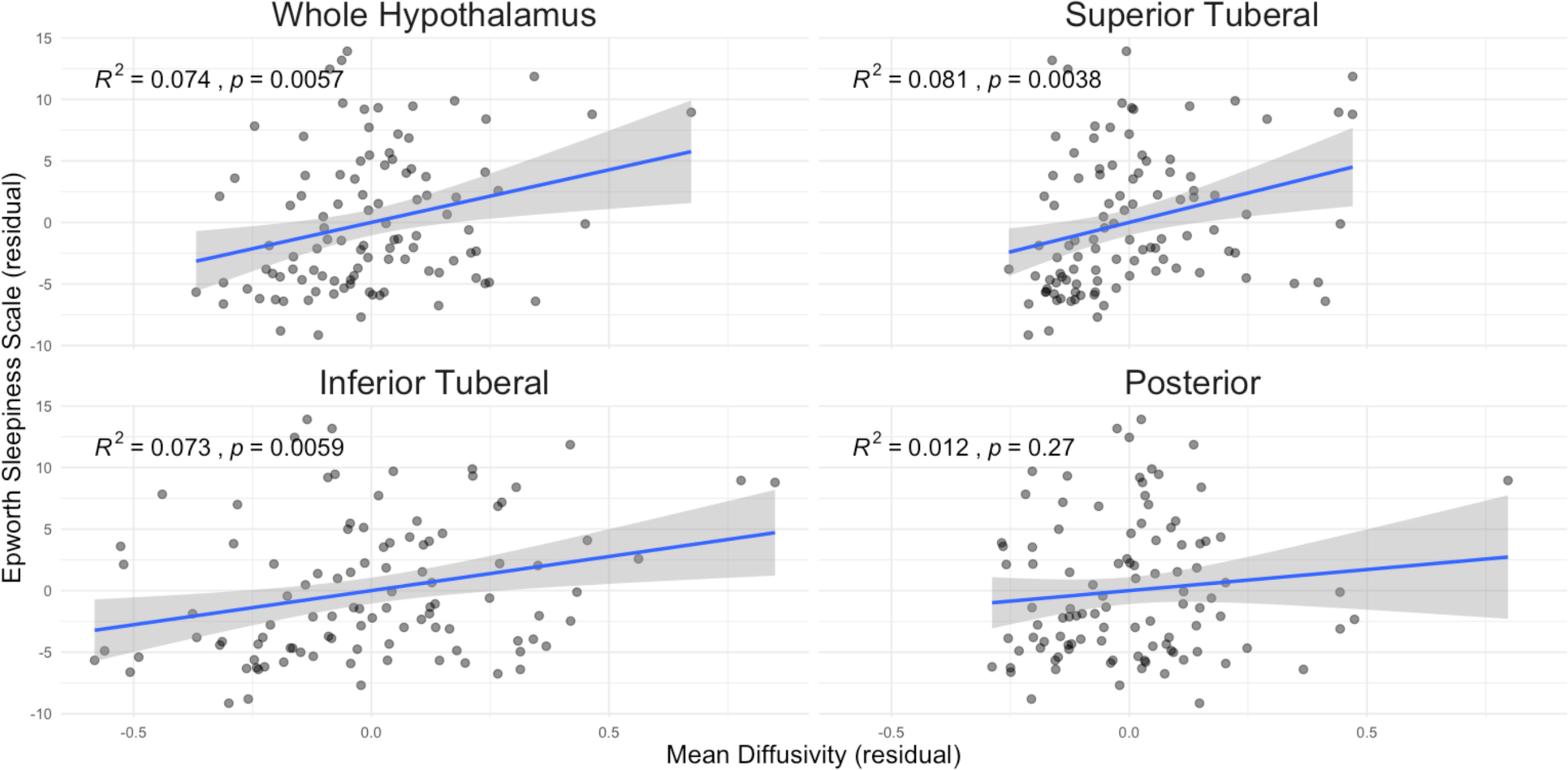
Hypothalamic mean diffusivity correlation with Epworth sleepiness scale scores. Plotted values are residualized Epworth sleepiness scores (y-axis) and hypothalamic MD (x-axis), adjusted for age and sex. The least squares best fit line is plotted in blue and the 95% confidence band shown in grey. Displayed p-values are not adjusted for multiple comparisons.

### Hypothalamic MD does not correlate with global cognition or motor scores

Next, we wondered if the association between hypothalamic MD and ESS reflected overall disease severity rather than a specific relationship with daytime sleepiness. To test this, we correlated hypothalamic MD with scores of global cognition (MMSE) and parkinsonian motor impairment (MDS-UPDRS Part III), covarying for age and sex. We found no correlation of MD with cognition or motor impairment (Figure 2).

**Figure 2:**
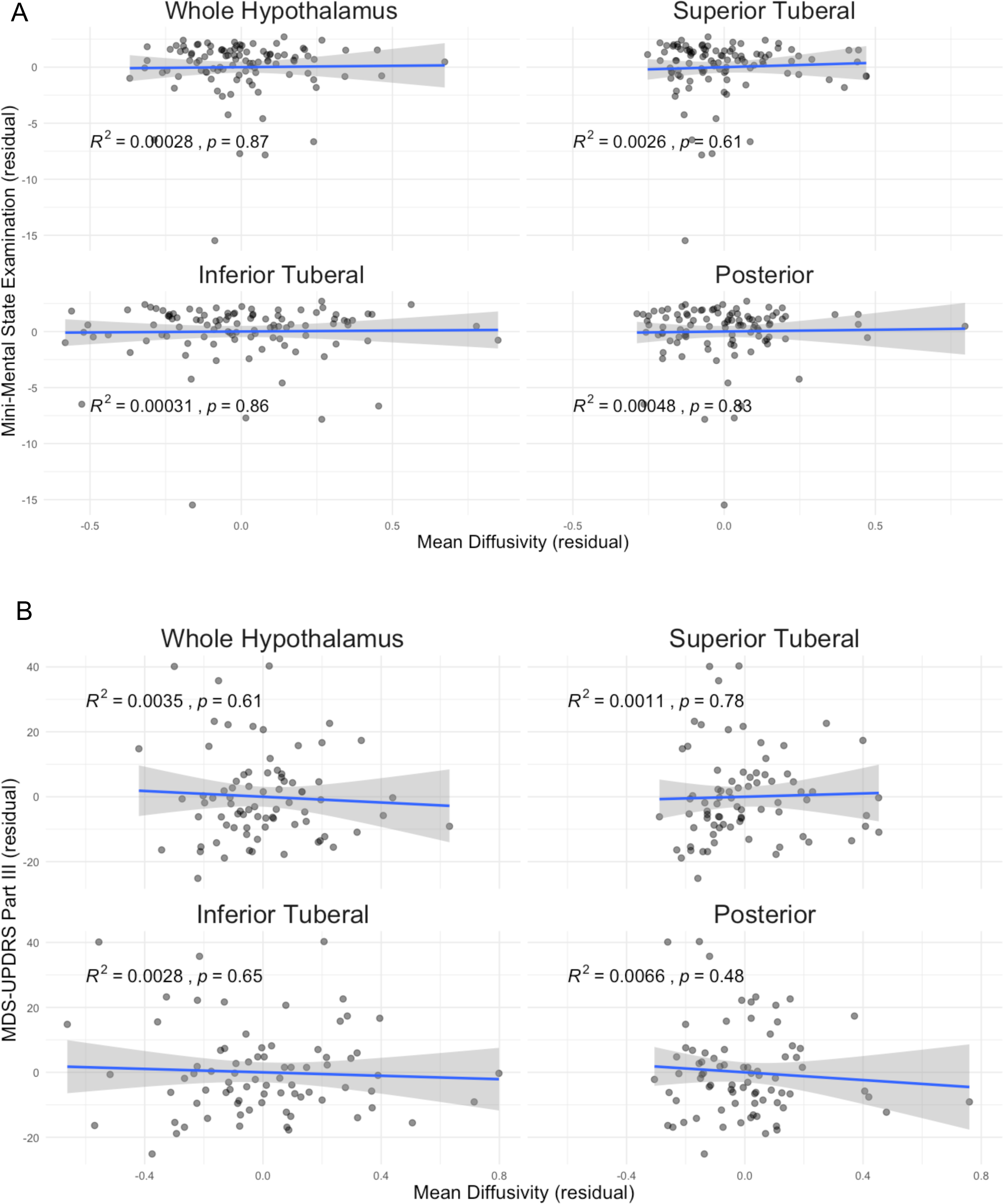
Hypothalamic mean diffusivity correlation with MMSE and MDS-UPDRS. Plotted values are residualized clinical scores (y-axis) and hypothalamic MD (x-axis), adjusted for age and sex. Results for global cognition are shown in (A) and motor impairment in (B). The least squares best fit line is plotted in blue and the 95% confidence band shown in grey. MMSE = Mini Mental State Exam; MDS-UPDRS III = Movement Disorder Society Unified Parkinson’s Disease Rating Scale Part III.

### Hypothalamic volume is not correlated with subjective sleepiness scores

Finally, we wondered to what extent the MD signal reflected hypothalamic atrophy. To test this, we correlated hypothalamic volumes with ESS, covarying for age and sex. We found no correlation of volumes with ESS in the whole hypothalamus nor any subregion (Figure 3).

**Figure 3:**
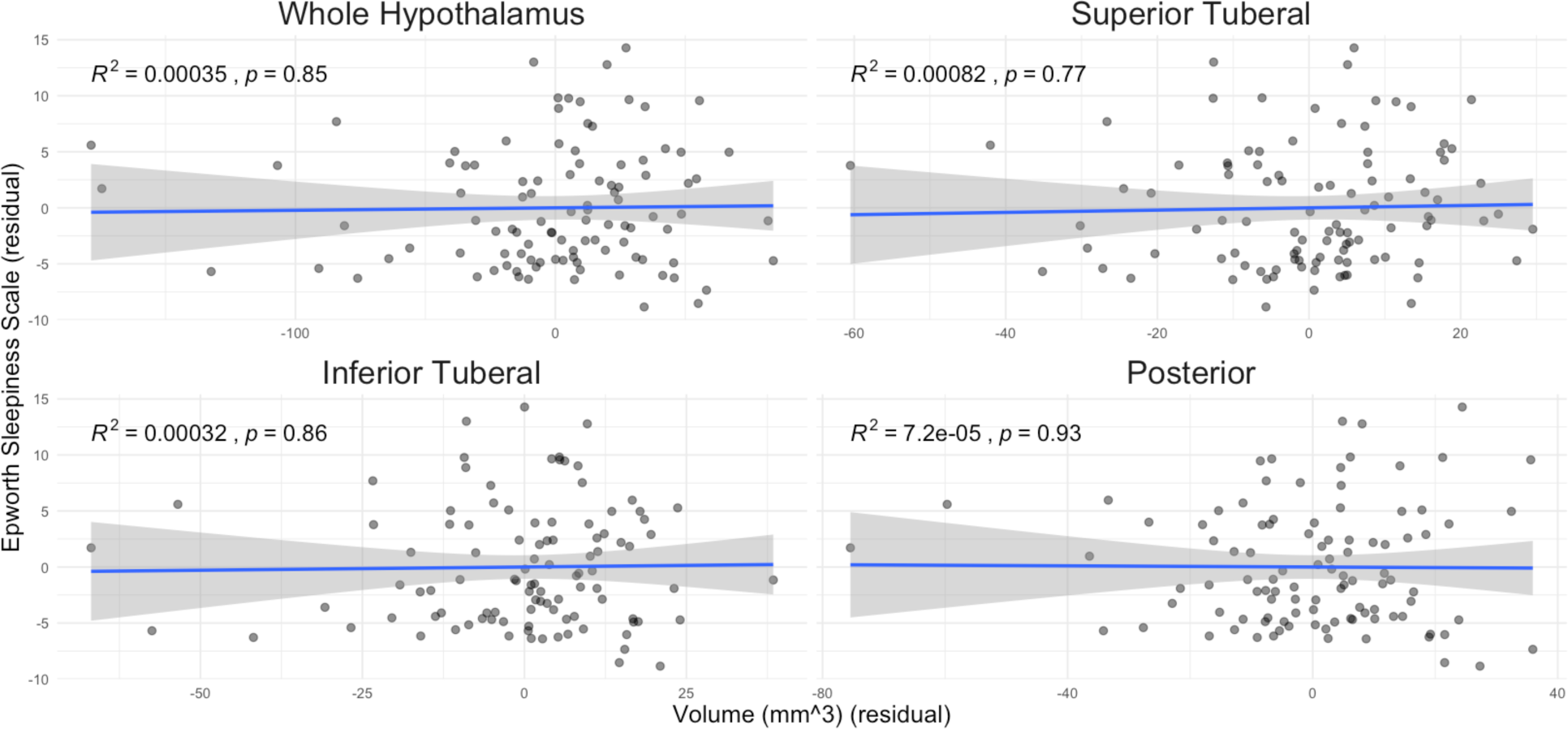
Hypothalamic volume correlation with Epworth sleepiness scale scores. Plotted values are residualized Epworth sleepiness scores (y-axis) and hypothalamic volume (x-axis), adjusted for age and sex. The least squares best fit line is plotted in blue and the 95% confidence band shown in grey. mm = millimeter

## Discussion

Excessive daytime sleepiness is a prevalent and debilitating symptom for individuals living with Lewy body disorders. The hypothalamus is the primary regulator of sleep-wake state and is known to be affected by Lewy body pathology. Here, we used diffusion MRI to show an association between microstructural changes in the hypothalamus and excessive daytime sleepiness in LBD. Specifically, we observed that increased mean diffusivity within the tuberal hypothalamus correlates with worse excessive daytime sleepiness. Furthermore, the MD changes do not appear to reflect global disease severity since they do not correlate with global cognition or parkinsonian motor signs. Notably, the diffusion signal changes are unlikely to reflect effects of atrophy since there was no correlation between ESS and hypothalamic volume.

Increases in mean diffusivity in grey matter generally reflect microstructural degeneration. The inferior tuberal subregion includes the TMN, which is involved by alpha-synuclein pathology by Parkinson’s Braak stage 3. The TMN is the sole source of histamine in the CNS, and histamine is a crucial mediator of arousal^48-50^. Thus, it is tempting to speculate that the observed radiologic signal reflects degeneration within the TMN with resultant sleepiness. Of course, this hypothesis must be evaluated by correlation with post-mortem tissue, which we intend to do among participants in our brain bank.

It has long been established that TMN neurons are involved by alpha-synuclein pathology^11,51,52^. However, despite the presence of Lewy bodies, associated neuronal loss is less clear^53^. While TMN neurons are lost in DLB^51^, they are preserved in PD^54,55^, with associated normal ^54,56^ or even increased ^57^ levels of histamine in the brain of PD patients at autopsy.

In the absence of neuronal loss, what might be the source of the mean diffusivity changes we observe? One possibility is that it reflects compensatory sprouting of histaminergic terminals in the setting of dopaminergic cell loss, as has been suggested previously in post-mortem studies^55,57^ and rodent models^58-60^. Of note, similar findings of compensatory TMN plasticity have been found in patients with narcolepsy, with increased histaminergic cell counts, nearly twice that of controls^61^. If histaminergic derangements contribute to EDS in LBD, this would point to potential targeted therapies, such as pitolisant, a histamine inverse agonist FDA-approved for treatment of hypersomnia in narcolepsy^62^.

Regarding MRI studies of the hypothalamus in LBD and its relation to sleep symptoms, there is limited data. Breen et al found that lower hypothalamic volume in PD correlated with decreased melatonin production^19^. This study also found decreased hypothalamic volume in PD (n=12) compared to controls (n=12). However, the volumetric difference was not reproduced in a larger study by Gorges et al (PD n=232, controls n=130)^20^. The latter study concluded that there was no atrophy in PD compared to controls, which is in line with postmortem findings of preserved neurons despite the presence of Lewy bodies in the hypothalamus. Furthermore, they found no correlation between hypothalamic volume and Epworth sleepiness scale, as in our cohort.

The automated hypothalamic segmentation approach used in our study^39^ differs from methods used in the above studies^19,20^. Breen et al. used a hypothalamic ROI from the WFU PickAtlas^63^, while Gorges et al. used an established manual method derived from histological analysis^64,65^. Billot et al.’s method was recently applied in an LBD cohort, where they similarly found no volumetric differences between clinical groups^21^.

### Strengths

The use of MD to assess the relationship between hypothalamic microstructure and daytime sleepiness in LBD is novel. While one other recent study analyzed hypothalamic microstructure in PD, associations with daytime sleepiness were not evaluated^21^. Our finding of a correlation between inferior tuberal MD and ESS suggest this subregion could be contributing to ESS symptoms and warrants further study. Another strength of our study is the use of an automated segmentation method, which improves the replicability and scalability of our analysis to other cohorts. Finally, detailed and standardized acquisition of clinical information in our cohort made it possible to assess for contributions/confounding by cognitive status and/or levodopa treatment side effects^66^.

### Limitations

While the overall size of the cohort was sufficient, there were very few participants with PDD or DLB (n=9 (8.8%)). Therefore, caution is warranted in generalizing the findings to beyond individuals with PD. Additionally, minor inaccuracies in segmentation of a small structure like the hypothalamus mean the measured MD differences could arise from a structure adjacent to the hypothalamus, such as the nucleus basalis of Meynert^37^. Relatedly, volume averaging effects resulting in contamination of the hypothalamus mask with CSF voxels from the adjacent third ventricle could skew MD. This should be worse in the presence of atrophy, and thus subtle volume loss may influence the observed MD. However, we found no relationship between volume and ESS. Additionally, while we control for age and sex, we did not control for presence of comorbid sleep disorders or use of sedative-hypnotic medications.

Finally, while we hypothesize that TMN degeneration could underly our findings, we must emphasize that our data cannot support such a specific claim. While MD could reflect neuronal loss, it is a nonspecific marker of tissue microstructure and can be influenced by astrogliosis^24^ and/or microglial activation^23,67^. Thus, we cannot be sure of the underlying process leading to MD signal differences. Furthermore, while we posit the specific involvement of the TMN, the inferior tuberal hypothalamic ROI contains multiple hypothalamic nuclei^39^ that could contribute to daytime sleepiness for example via altered energy homeostasis (ventromedial nucleus)^68^.

## Conclusion

Increased MD in the hypothalamic inferior tuberal subregion correlates with sleepiness but not cognition or motor symptoms in LBD. This suggests that pathology within this region, which includes the wake-promoting tuberomammillary nucleus, could contribute to EDS symptoms. Future work will correlate *in vivo* MRI with post-mortem histopathology to clarify the anatomic basis of the observed microstructural changes. If sleepiness symptoms relate to histaminergic degeneration, histamine-stimulating therapies could be beneficial in this population.

## Acknowledgments

None

